# Are we ready when COVID-19 vaccine is available? Study on nurses’ vaccine hesitancy in Hong Kong

**DOI:** 10.1101/2020.07.17.20156026

**Authors:** Kin On Kwok, Kin Kit Li, Wan In Wei, Arthur Tang, Samuel Yeung Shan Wong, Shui Shan Lee

**Affiliations:** JC School of Public Health and Primary Care, The Chinese University of Hong Kong, Hong Kong Special Administrative Region, China; Stanley Ho Centre for Emerging Infectious Diseases, The Chinese University of Hong Kong, Hong Kong Special Administrative Region, China; Shenzhen Research Institute of the Chinese University of Hong Kong, Shenzhen, China; Department of Social and Behavioural Sciences, City University of Hong Kong, China; Department of Software, Sungkyunkwan University, Seoul, South Korea

## Abstract

**Introduction:** Nurses are considered a trustworthy source of vaccine-related information to build public confidence in vaccination. This study estimated nurses’ influenza vaccine uptake and intention to receive COVID-19 vaccine when available, and examined the corresponding psychological antecedents.

**Methods:** A cross-sectional online survey among nurses was conducted during the main COVID-19 outbreak in Hong Kong between mid-March and late April 2020. Demographics, influenza vaccination, intention to have COVID-19 vaccine, the 5C vaccine hesitancy components (i.e., confidence, complacency, constraints, calculation, and collective responsibility), work stress and COVID-related work demands (i.e., insufficient supply of personal protective equipment, involvement in isolation rooms, and unfavorable attitudes towards workplace infection control policies) were reported.

**Results:** The influenza vaccination coverage and the proportion intending to take COVID-19 vaccine were 49% and 63%, respectively, among 1205 eligible nurses. Influenza vaccine uptake was associated with working in public hospitals and all 5C constructs, whereas stronger COVID-19 vaccination intention was associated with younger age, more confidence, less complacency and more collective responsibility towards the vaccine. COVID-19-related demands were associated with greater work stress, and hence stronger COVID-19 vaccination intention.

**Conclusion:** Vaccine uptake/intention was well predicted by the 5C constructs. With less work stress among nurses in the post-pandemic period, the intention to take COVID-19 vaccine will likely drop. The 5C constructs should be infused in vaccination campaigns. While a COVID-19 vaccine could be ready soon, communities are not ready to accept it. More research work is needed to boost the uptake.

## INTRODUCTION

Vaccination is considered an effective approach to prevent infection and substantially reduce the mortality of many infectious diseases such as influenza and HPV^12^. However, vaccine hesitancy, a behaviour with delay in acceptance or refusal of vaccines despite availability of vaccination services, has depolarized the vaccine-supporters and their anti-vaccine counterparts. The World Health Organization (WHO) considered this phenomenon as one of the ten threats in global health in 2019.Three main factors are contributing to vaccine hesitancy: (i) individuals may lack confidence in and be fearful towards vaccines, especially with the misunderstanding that vaccines pose a risk of infection; (ii) individuals do not perceive a need for a vaccine or do not value the vaccine. For example, the disease severity may be underestimated that individuals hold a complacent attitude towards the need for prophylaxis; and (iii) it may be difficult to access the vaccine^3^. This phenomenon was most conspicuous in the uptake decision for influenza vaccine in the general population. Ten years since the influenza pandemic in 2009, about half of the population in the United States did not have a seasonal influenza vaccine in 2019^4^.

During the epidemic, Health Care Workers (HCWs) in the hospital can be considered as one of the high-risk groups. HCWs’ infection risk could be amplified during the ongoing epidemic due to various factors including continuous exposure to patients, shortages of personal protective measure (PPE) supply and inadequate infection control training after immediate response to outbreak among HCWs. During the 2003 severe acute respiratory syndrome (SARS) epidemic in Hong Kong, the first large transmission cluster occurred in Prince of Wales Hospital where HCWs accounted for a substantial proportion of infection with 43.6% among cases admitted to this hospital^56^. HCWs were consequently responsible for about a quarter of total SARS infections in Hong Kong^7^. While we were noted for SARS-coronavirus (SARS-CoV) in the 2003 epidemic, another five human coronaviruses (OC43, 229E, NL63, HKU1, Middle East Respiratory Syndrome-CoV) had been circulating for decades among human populations^8^. With their large genetic diversity and frequent genome recombination, a new emerging coronavirus-SARS-CoV-2 was first identified in Wuhan, China in late December 2019, and resulted in subsequent Coronavirus COVID-19 infections in other provinces in mainland China, Hong Kong and ultimately all over the world. As of 24 February 2020, 3387 HCWs in medical facilities of 77262 COVID-19 laboratory confirmed cases in China (4.4%) were recorded, of which 23 died from the infection^9^. With the challenge of possible resurgence of COVID-19 in the near future, increasing the proportion of immune individuals among HCWs and the general population to the disease by vaccination as the indirect protection would be the only viable option to avoid nosocomial infection. A survey conducted during the period of nationwide lockdown in France showed that a quarter of the adult population (26%) refused to vaccinate against SARS-CoV-2 when it becomes available and were skeptical about its effectiveness^10^.

Understanding HCWs’ vaccine hesitancy has substantial implications on public health administrations during epidemics. HCWs’ infections take a direct bow on the healthcare system during epidemics in terms of reducing available healthcare workforce. They are usually at the front end of fighting with epidemics, and some of them are required to routinely perform procedures with high risks of contracting with pathogens. HCWs also serve as potential vectors for nosocomial transmission that protection them from epidemics play a pivotal role in infection control. Besides, HCWs were considered as a trustworthy source of vaccine-related information for patients^11^. The WHO vaccine advisory group also highlighted that their role in building public confidence in vaccines^12^. They convey the message of vaccination benefits and address the worries and concerns of the patients on a new developed vaccine. However, the influenza vaccine coverage among nurses was only slightly greater than 30%^13^. In light of this, improving our understanding of determinants favoring vaccine uptake among nurses, a core group of HCWs, could have broader policy implications for future COVID-19 vaccine acceptability and dissemination.

In this study, we first estimated the proportion of nurses with the intention in taking COVID-19 and influenza vaccine uptake coverage. Second, a comparison analysis was conducted to examine how well the vaccine hesitancy domains can predict both vaccines uptake decisions. Third, the association of work stress with vaccine uptake decisions during the COVID-19 pandemic and factors associated with the stress were also investigated.

## METHOD

A cross-sectional online self-administered survey was conducted among nurses in Hong Kong. The survey collected items including demographics (year of birth, sex, rank, presence of chronic diseases), level of contacts with patients, influenza vaccine uptake and intention to take COVID-19 vaccine when available, statements measuring the 5 domains of vaccine hesitancy (see details below), work stress level, supply of PPE, involvement in isolation rooms, and attitudes towards workplace infection control policies.

This study was approved by the Survey and Behaviour Research Ethics Committee of The Chinese University of Hong Kong (reference number: SBRE-19-251).

### Participants

The required sample size to estimate seasonal influenza vaccine coverage and COVID-19 vaccine intention was 1049 based on an estimated population of 60000 registered or enrolled nurses in Hong Kong, a 3% margin of error, a 95% confidence interval, and a prevalence rate at 50%. To account for a 30% loss from invalid cases (ineligible or incomplete cases), the sample size required was 1499. The online survey was disabled when the sample size was achieved.

In collaboration with the Association of Hong Kong Nursing Staff (AHKNS), registered nurses, enrolled nurses, nursing students and trainees working in public or private medical facilities were recruited in this study in the period from 16 March 2020 to 29 April 29 2020. Among over 50000 registered or enrolled nurses in Hong Kong, over 60% were members of AHKNS. A sample recruited via AHKNS would be rather representative of the nurses in Hong Kong. Participants were compensated with a coupon of HKD 25. Nursing students and retired nurses were excluded from this analysis.

### Measures

Vaccine hesitancy was measured using a 15-item tool developed from a “5C model” of psychological antecedents to vaccination^14^. Each of the 5 antecedents including confidence, complacency, constraints, calculation, and collective responsibility, was assessed by 3 rating items on a 7-point scale (1=*strongly disagree*; 7=*strongly agree*). Mean scores of items under each domain were computed, with higher average score indicating stronger agreement of the corresponding domain.

Work stress was measured by a single item asking participants to self-rate their level of work stress after the outbreak of COVID-19 on an 11-point scale (0=*no stress at all*; 10=*the maximum stress*). Insufficient supply of PPE was measured by asking participants to report any shortage of 7 PPE and an open option (1=*yes*; 0=*no*). The higher the total score, the more insufficient supply of PPE was. A single item asking participants whether their job duties included work in infection isolation rooms (1=*yes*; 0=*no*). Attitudes towards workplace infection control policies were measured by 3 items stating if the workplace infection control policies were timely, sufficient, and effective, respectively, on a 5-point rating scale (1=*strongly disagree*; 5=*strongly agree*).

Seasonal influenza vaccine uptake was measured by self-reported vaccination in 2019/20 while COVID-vaccine uptake intention was measured by a single item asking participants how likely they will take the COVID-19 vaccine when available on a 11-point likert scale (0=*definitely no*; 10=*definitely yes*).

### Statistical Analysis

To examine the potential bias on excluded cases, the sample characteristics were compared between those with excluded and analyzed responses. To further examine the sample representativeness, a couple of sample characteristics were compared with those reported in two large-scale Health Manpower Surveys conducted by the Department of Health of the Hong Kong SAR government. We summarized the characteristics of the study participants with descriptive statistics such as mean, frequency, percentage and 95% binomial confidence interval (bCI) and their bivariate correlations. A factor analysis using principal axis factoring approach was conducted to examine the factorial validity of the 5C model in the current population. Multivariate linear and logistic regression models were applied to identify factors associated with COVID-19 vaccine uptake intention and influenza vaccine uptake decision respectively. The mediating effect was also tested using path analysis with 2000 bootstrapped samples. A statistical significance was based on p-value of 0.05. All analyses were conducted in R (v3.6.3) and Stata 16.0.

## RESULTS

A total of 1660 attempts to complete the survey were recorded, of which 1205 respondents were retained for the analyses. Excluded cases were those who had retired (*n*=37) or full-time nursing students (*n*=95), or provided incomplete responses (*n*=323). No statistically significant difference was found in sex composition, presence of chronic diseases, being a AHKNS member and ever had influenza vaccination between those from complete and incomplete responses. Those with complete responses, however, were more likely to be degree holders, working in the public service run by Hospital Authority, older, have more frequent contact with patients, and a weaker intention to take COVID-19 vaccine than those with incomplete responses. Registered nurses (80%) were slightly overrepresented in our sample as compared with the percentage of registered nurses in the Nursing Council of Hong Kong (75%), χ^2^(1)=8.62, *p*=.003. Nurses in this sample were slightly more likely to be women, degree holders, less likely to work in Hospital Authority, and younger as compared with those in the Health Manpower Survey, χ^2^(1)=6.89, *p*=.009. (**Table S1)**

**Table 1** shows sample characteristics and their bivariate associations with influenza vaccine uptake and COVID-19 vaccine intention, respectively. The mean age of the sample was 40.79 years (*SD*=10.47). Most participants were female (90%) and AHKNS members (96%). More than half of the participants worked in the public hospitals (56%). Participants reported high exposure to patients (*M*=4.35 on a scale of 1-5; *SD*=1.23).

**Table 1.** Sample Characteristics, Crude Odds Ratios Predicting Influenza Vaccination, and Correlations with COVID-19 Vaccine Intention (N = 1205)

| Predictor (range) | Mean / % | SD | Influenza vaccination | COVID-19 vaccination |
| --- | --- | --- | --- | --- |
|  |  |  | OR (95% CI) | intention<br>*r* |
| Age (21-71) | 40.79 | 10.47 | **1.01 (1.00, 1.02)** | -.03 |
| Sex (1 = women) | 89.71% |  | 0.98 (0.68, 1.42) | -.02 |
| Chronic diseases (1 = yes) | 12.70% |  | **1.54 (1.09, 2.17)** | .00 |
| Public hospitals (1 = yes) | 56.35% |  | 1.25 (1.00, 1.57) | -.03 |
| Patient contact frequency (1-5) | 4.23 | 1.24 | 0.98 (0.89, 1.07) | .01 |
| Confidence (1-7) | 4.94 | 1.21 | **3.25 (2.81, 3.77)** | .38*** |
| Complacency (1-7) | 3.64 | 1.24 | **0.56 (0.51, 0.62)** | -.20*** |
| Constraints (1-7) | 3.15 | 1.28 | **0.69 (0.63, 0.76)** | -.06* |
| Calculation (1-7) | 5.61 | 0.88 | 1.03 (0.90, 1.17) | .11*** |
| Collective responsibility (1-7) | 5.28 | 1.16 | **2.43 (2.13, 2.78)** | .33*** |
| Work stress (0-10) | 7.38 | 2.06 | **1.06 (1.00, 1.12)** | .21*** |
| Insufficient supply of PPE (0-8) | 2.79 | 1.87 | 0.96 (0.91, 1.02) | .04 |
| Involvement in isolated rooms (1 = yes) | 32.70% |  | 0.98 (0.77, 1.25) | -.03 |
| Attitudes toward control policies (1-5) | 2.56 | 1.05 | 1.14 (1.02, 1.27) | -.03 |

\* $p < .05$ , \*\*\* $p < .001$ .
PPE: personal protective equipment.
Significant odds ratios (95% confidence interval) are presented in bold face.

The influenza vaccine coverage in the 2019-20 winter season was 49% (95% bCI: 47%, 52%). Univariate associated determinants with higher influenza vaccine uptake were older age, presence of chronic diseases, stronger vaccine confidence, collective responsibility, and work stress; and weaker vaccine complacency and constraints. Intention to take COVID-19 vaccine when available was 6.52 (on a scale of 0-10; *SD*=2.83), which could be translated to 63% (95% bCI: 60%, 66%) reporting they were likely to vaccinate (scored 6 or above). Univariate factors associated with stronger intention to take COVID-19 vaccine were stronger vaccine confidence, calculation, collective responsibility, and work stress; and weaker complacency and constraints. Correlations among the studied variables and Cronbach’s alpha coefficients for composite measures are presented in **Table S2**.

The results of a parallel analysis showed that 5 factors should be retained for the vaccine hesitancy measure (**Table S3**). Bartlett’s test, χ^2^(105)=7841.71, *p*<.001, and KMO measure (.82) also supported the factorability and sufficiency of the data. Using Oblimin rotation, all items conformed to the original factor structure, with factor loadings ranging from .63 to .84, except the only reverse item tapping collective responsibility. It was removed and subsequently increased the Cronbach’s alpha coefficient of collective responsibility from 0.62 to 0.82.

To explore the relationship between the progression of number of daily confirmed cases and nurses’ intention to take COVID-19 vaccine, we overlaid the averaged intention of each reporting day over the epidemic curve of Hong Kong (**Figure S1**). The data collection period covered the main wave of COVID-19 outbreak in Hong Kong. The data reflected that intention was high and stable during the burst of imported cases and local transmissions. A sudden drop in the intention to take COVID-19 vaccine was observed when the number of confirmed cases dropped at the end of the main wave of the outbreak. The level of intention was reinstated and less stable afterward.

### Validity of 5C model in influenza vaccine uptake and COVID-19 vaccine intention

The reference models to predict influenza vaccine uptake or COVID-19 vaccination intention included only covariates. Adding 5C into the influenza vaccination model increased the pseudo *R*^2^ from 0.71% to 29.91%. In the final logistic regression model, influenza vaccination was associated with working in public hospitals, *aOR*=1.56 (95%CI=1.16, 2.10), and having stronger vaccine confidence, *aOR*=2.70 (2.27, 3.22), and collective responsibility, *aOR*=1.67 (1.40, 1.98), and weaker complacency, *aOR*=0.69 (0.60, 0.79), constraints, *aOR*=0.83 (0.73, 0.94), and calculation, *aOR*=0.62 (0.51, 0.75). In comparison, adding 5C into COVID-19 vaccination model increased the *R*^2^ from 0.27% to 17.70%. In the final multiple regression model, intention to take COVID-19 vaccine was associated with being younger, *β*=-.07, *p*=.02, and having stronger vaccine confidence, *β*=.29, *p*<.001, and collective responsibility, *β*=.12, *p*<.001, and weaker complacency, *β*=-.11, *p*<.001. **Table 2** shows the coefficients of the two regression models. When COVID-19 vaccination intention was dichotomized as likely (score 6-10) and not likely (score 0-5), the pseudo *R*^2^ of the model was 10.19%. The coefficients of the dichotomized COVID-19 vaccination intention model are presented in **Table S4**.

**Table 2.**
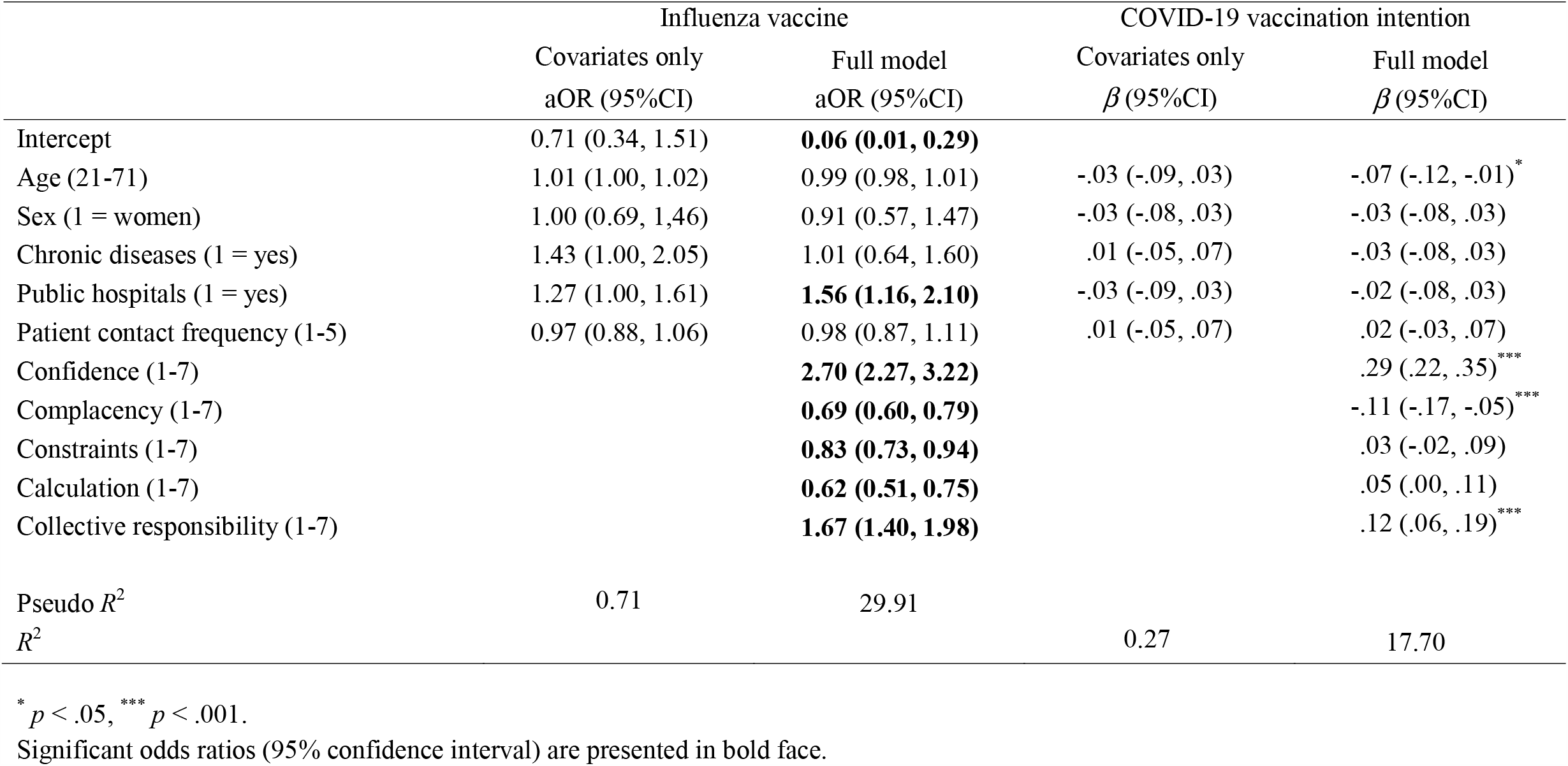
Effects of the 5C Model of Vaccine Hesitancy on Influenza Vaccination and COVID-19 Vaccination Intention

### Effects of COVID-19 demands on vaccination intention via work stress

To assess whether work stress mediated the association between COVID-19-related demands and vaccination intention, we conducted a path analysis with 2000 bootstrapped samples (**Figure 1, Table S5**). The indirect effects of insufficient supply of PPE, *β*=.04, *p*<.001, involvement in isolation rooms, *β*=.09, *p*=.005, and attitudes towards control policies of public authorities, *β*=-.07, *p*=.001, on COVID-19 vaccination intention via work stress were significant. Insufficient supply of PPE, *β*=.17, *p*<.001, involvement in isolated rooms, *β*=.39, *p*=.001, and unfavorable attitudes towards control policies of public authorities, β=-.29, *p*<.001, were associated with work stress. Work stress was subsequently associated with vaccination intention, *β*=.22, *p*<.001, controlling for the predictors in the previous COVID-19 vaccination intention model and the influenza vaccination status.

**Figure 1.**
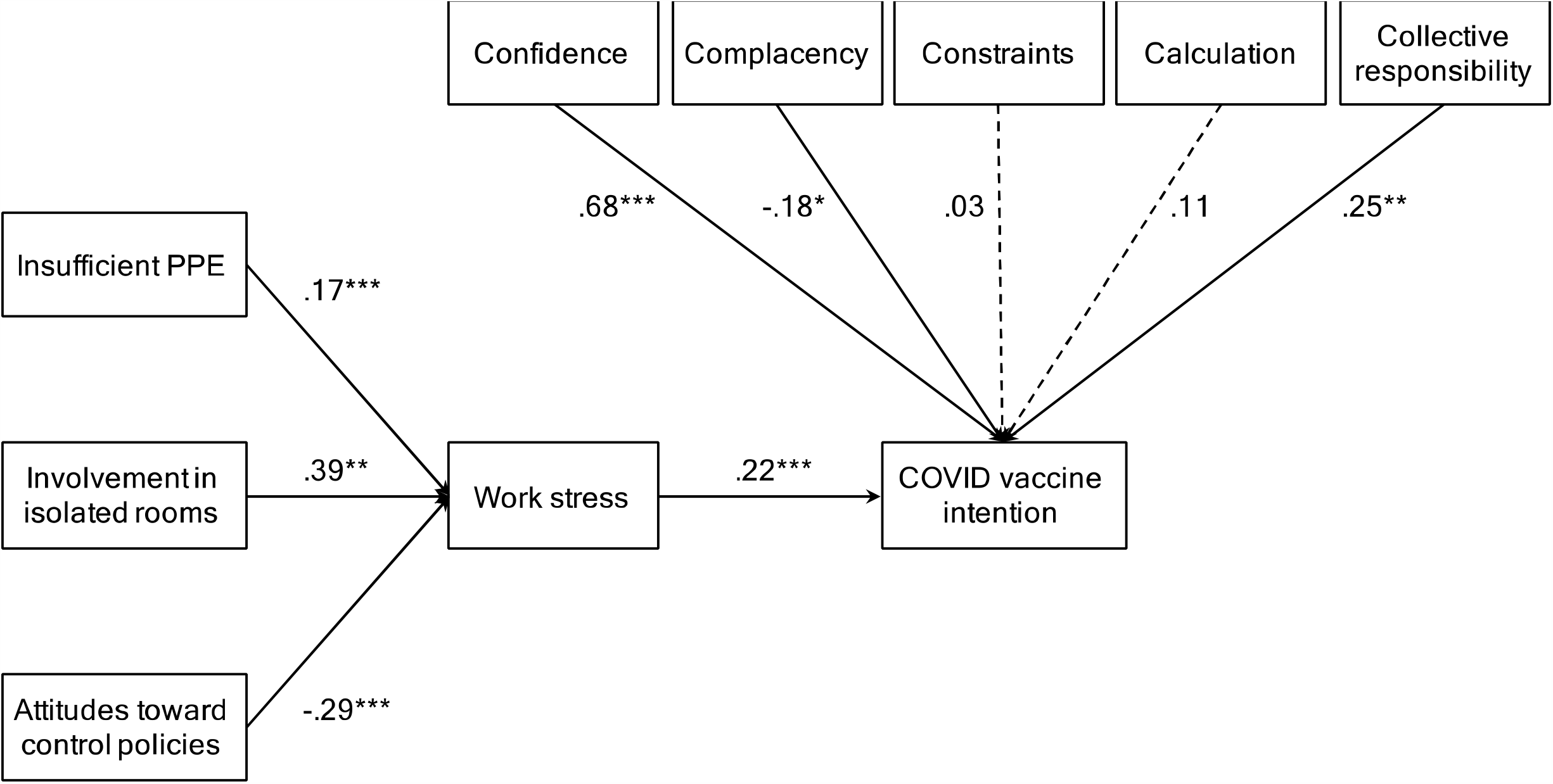
The effects of 5C and the mediation effect of work stress on COVID-19 vaccination intention.

## DISCUSSION

With the large-scale cross-sectional online survey with more than 1000 respondents during the main wave of outbreak in the early phase of COVID-19 epidemic, this was the first study presenting the uptake behaviour/intention of both influenza and potential COVID-19 vaccine among nurses in Hong Kong. We report the estimates of both influenza vaccine uptake and intention to have COVID-19 vaccine and identify their associated factors.

Despite the uncertainty of vaccine attributes such as effectiveness, side effects and duration of protection, more than half of the respondents (63%) indicated that they were likely to opt for COVID-19 vaccine when it becomes available. Younger age, stronger confidence and collective responsibility, and weaker complacency were associated with stronger intention to be vaccinated. Also, COVID-19-related demands including insufficient supply of PPE, involvement in isolated rooms and poorer attitudes towards workplace control policies among nurses in the early phase of the epidemic in Hong Kong were associated with greater work stress which in turn resulted in stronger intention to have COVID-19 vaccine. Similar to a risk perception survey among the general population of Hong Kong^15^, the experience of SARS contributing to strong psychological responses, as reflected in nurses’ pressure level, underlined their vaccine uptake intention for COVID-19. This also applies to influenza vaccination. About half of the respondents (49%) reported to receive influenza vaccine in the 2019/2020 season. This estimate is statistically higher than those observed in similar surveys from the same population in 2013/14, 2014/15, 2015/16, 2016/17 seasons (32%, 28%, 33% and 36% respectively)^13,16,17^. This high rate may possibly be due to the similarity of COVID-19 symptoms with those observed in influenza or other respiratory diseases^18^. Working in the public hospital, more confidence, less complacency, less constraints, less calculation and more collective responsibility were associated with the decision to have influenza vaccine uptake. The 5C model was more predictive of influenza vaccine uptake than intention to take COVID-19 vaccine based on the pseudo *R*^2^ coefficients of the models.

Our study has several important public health implications. ***First***, identification of determinants associated with COVID-19 vaccine uptake intention and influenza vaccine uptake decision helps inform future vaccination campaigns. Older nurses with less intention to have COVID-19 vaccine may contribute to possible nosocomial infection by their close contact with COVID-19 patients in the hospital. As older individuals are more susceptible to COVID-19 than younger individuals^19^, in the absence of vaccine uptake, they will very likely be a high-risk group in the next wave of COVID-19 epidemic. Older and experienced nurses are particularly valuable in public health emergency. The protection to this high-risk and highly valuable subgroup is particularly important during an outbreak. With sporadic cases or fewer imported cases after the major epidemic globally and further improvement in infection control practices, nurses will likely have relatively lower work stress. In this connection, they may be less likely to have intention to take COVID-19 vaccine when the vaccine is available. Another challenge is that age is a mortality risk factor for COVID-19 infection, but older nurses are less likely to take the vaccine. With their experience, they are likely to be the role models of the junior nurses. Health authorities should tailor a vaccination program to nurses, in particular older nurses, to have COVID-19 vaccination. Future research is also needed in order to investigate why older nurses have a higher vaccination hesitancy, and explore potential strategies in consciousness raising and attitude changing towards vaccination.

***Second***, uptake of the safe and effective vaccine could only be considered as an additional measure to help control the COVID-19 pandemic. Assuming the population of COVID-19 vaccine coverage is similar to that observed among nurses in this study with a conservative effectiveness of 50%, the spread of infection will be halted if the effective reproductive number Rt, a measure to estimate the number of secondary cases generated by an index case in the presence of interventions, is below 1.45. Apart from vaccination campaigns to boost uptake rate and continuous development of antiviral therapy, the health authority should further consider to conduct the modelling studies to explore the optimal levels of assorted interventions including encouragement of social-distancing adoption, border controls, active case surveillance and contact tracing to maintain the epidemic in a manageable level.

***Third***, more emphasis should be put on psychological components when implementing the national-wide vaccination program. Our statistical framework suggested that the variation of psychological constructs in the 5C model contributed a significant proportion to explain both influenza vaccine uptake and COVID-19 vaccine uptake intention. Our findings were consistent with a previous study that the 5C vaccine hesitancy scale could well examine the psychological antecedents of influenza vaccination^14^. However, the power of 5C was weaker in predicting COVID-19 vaccine uptake intention. It is not surprising that calculation and constraints in the 5C hesitancy model were found to be not associated with this intention. Given very limited information related to COVID-19 vaccine during the early phase of the epidemic, respondents were not able to perform an extensive information search and evaluate their synonyms for the possible perceived barriers on the new vaccine. The validity of the 5C model may increase as there is more information about the new vaccine. Further validation work of vaccine hesitancy models on COVID-19 vaccine is warranted. When the strongest interventions such as mandatory vaccination or opt-out policies^20^ are not ethically justified, targeting the 5C components through evidence-informed interventions^21^, health communication approaches^22^, and new media^23^ may be some viable options.

This study has a couple of limitations. ***First***, a convenience sampling approach may result in potentially biased estimates. ***Second***, this was a cross-sectional study which could not infer the causal relationship. ***Third***, possible recall bias may occur in self-reporting measurements. ***Fourth***, not all components in the 5C vaccine hesitancy model could address the intention to have COVID-19 vaccine hesitancy when the vaccine attributes are not available. ***Fifth***, the intention to receive COVID-19 vaccine may be sensitive to the time-varying infection and mortality rate of the ongoing pandemic.

## CONCLUSION

This study provided additional validity evidence for the 5C vaccine hesitancy model and showed its potential in predicting and promoting COVID-19 vaccine when available. While we are cautiously optimistic that the vaccine will decrease the transmission, its ability to control the pandemic is dependent on multiple factors such as uptake rate and vaccine effectiveness. If only 63% of nurses during the main outbreak in Hong Kong intended to take the COVID-19 vaccine, we anticipate that promoting the vaccine to the general public in the post-pandemic period will be much more challenging. While a vaccine could be ready in a few months, our community and many alike are not ready to accept it. More research work is needed to optimize the uptake of the vaccine, our best hope so far.

## Data Availability

The anonymous dataset to generate this research will be available upon request.

## ACKNOWLEDGEMENT

We thank the Association of Hong Kong Nursing Staff and Li Ka Shing Institute of Health Sciences, The Chinese University of Hong Kong, for technical support. We also thank Mandy Li Man Wai for administrative support. We acknowledge Research Fund for the Control of Infectious Diseases, Hong Kong (Number: INF-CUHK-1), General Research Fund (Number: 14112818), Health and Medical Research Fund (Ref: 18170312) and Wellcome Trust (UK, 200861/Z/16/Z).

## EXCLUSIVE LICENCE

This manuscript is published under the Creative commons licence (CC BY).

## CONTRIBUTORSHIP

Kin On KWOK (KOK), Kin Kit LI (KKL), Samuel Yeung Shan WONG (SYSW) and Shui Shan LEE (SSL) conceptualized the study; KOK and KKL analysed the data; KOK and KKL wrote up the first draft of the manuscript; Wan In WEI (WIW), Arthur TANG (AT), SYSW and SSL reviewed and edited the manuscript; SSL acquired funding and performed project administration.

